# A Cost Analysis of Childbirth for Pregnant Women with COVID-19 in Nigeria’s Epicentre

**DOI:** 10.1101/2020.09.22.20199265

**Authors:** Aduragbemi Banke-Thomas, Christian Chigozie Makwe, Mobolanle Balogun, Bosede Bukola Afolabi, Theresa Amaogechukwu Alex-Nwangwu, Charles Anawo Ameh

## Abstract

The Coronavirus disease 2019 (COVID-19) has been a major disruptor of health systems globally. Its emergence has warranted the need to reorganize maternity services for childbirth. However, it is not known if this comes at an additional cost to women. We conducted a hospital-based cost analysis to estimate the out-of-pocket cost of spontaneous vaginal delivery (SVD) and caesarean delivery (CD). Specifically, we collected facility-based and household costs of all nine pregnant women with COVID-19 who were managed between 1^st^ April and 30^th^ August 2020 at the largest teaching hospital in Lagos, the epicentre of COVID-19 in Nigeria. We compared the mean facility-based costs for the cohort with costs paid by pregnant women pre-COVID-19, identifying major cost drivers. We also estimated what would have been paid without subsidies, testing assumptions with a sensitivity analysis. Findings showed that total utilization cost ranged from US$494 (N190,150) for SVD with mild COVID-19 to US$4,553 (N1,751,165) for emergency CD with severe COVID-19. Though 32-66% of facility-based cost has been subsidized, cost of SVD and CD have doubled and tripled respectively during the pandemic compared to those paid pre-COVID. Out of the facility-based costs paid, cost of personal protective equipment (PPE) was the major cost driver (50%) for SVD and CD. Supplemental oxygen was a major cost driver when women had severe COVID-19 symptoms and required long admission (48%). Excluding treatment costs specifically for COVID-19, mean facility-based costs for SVD and CD are US$228 (N87,750) and US$948 (N364,551) respectively. Our study demonstrates that despite cost exemptions and donations, utilization costs remain prohibitive. Regulation of the PPE and medical oxygen supply chain can help drive down utilization cost and reduce mark-ups being passed to users. The pandemic offers an opportunity to expand advocacy for subscription to health insurance schemes in order to avoid any catastrophic health expenditure.

**KEY MESSAGES:**

- Total utilization cost ranged from US$494 (N190,150) for spontaneous vaginal delivery with mild COVID-19 to US$4,553 (N1,751,165) for emergency caesarean delivery with severe COVID-19.
- Cost of personal protective equipment was the major cost driver (50%) for vaginal and elective caesarean deliveries. Medical oxygen was a major cost driver when women had severe COVID-19 symptoms (48%) and required long admission.
- Though 32-66% of total cost have been subsidized, facility-based cost of vaginal and caesarean deliveries has doubled and tripled respectively during the pandemic compared to those paid pre-COVID.
- The study findings highlight the urgent need to implement strategies that can help to minimize the rising costs that pregnant women with COVID-19 face in accessing and utilizing critical intra-partum care.

## Introduction

The Coronavirus disease 2019 (COVID-19) is caused by a novel strain of coronavirus known as severe acute respiratory syndrome coronavirus 2 (SARS-CoV-2). Following its emergence in Wuhan, China in December 2019, the disease was declared a global pandemic by the World Health Organization (WHO) on 11^th^ March 2020 and has since become a major disruptor of health systems and life in general (WHO, 2020a, 2020b). As of September 2020, there have been over 24 million confirmed cases, including almost 850,000 deaths globally (WHO, 2020b). This has come on the heels of significant gains in global maternal mortality where there has been a 50% reduction in deaths related to pregnancy and childbirth over the past two decades. In 2017, the WHO estimated that there were 295,000 maternal deaths worldwide (WHO *et al*., 2019). However, modelled estimates from a study published early on in the COVID-19 pandemic predicted that between 8·3 and 38·6% increase in maternal deaths should be expected per month across 118 countries (Roberton *et al*., 2020). Such increments do not bring countries any closer to achieving the global consensus target for the next decade of reducing maternal mortality ratio to 70 per 100,000 live births (United Nations, 2016).

Nigeria accounts for 25% of the global maternal deaths, with 512 maternal deaths reported per 100,000 live births (National Population Commission and ICF International, 2019; WHO *et al*., 2019). Approximately 60% of maternal deaths in the country occur either during delivery (intrapartum) or six-week period after delivery (post-partum), with the leading causes of deaths being bleeding, high blood pressure, and infection in pregnancy (Merdad and Ali, 2018). Access to skilled health personnel to provide skilled birth attendance and emergency obstetric care (EmOC) have been shown to reduce maternal mortality by as much as 50% and stillbirths by between 45 and 75% (Paxton *et al*., 2005). However, while multiple factors contribute to limiting access to these essential maternity services, one key barrier that has been widely documented is the cost of service utilization (WHO *et al*., 2019). In Africa, almost all women (97%) are delivered by spontaneous vaginal delivery (SVD) or caesarean delivery (CD), which can be elective (planned) or emergency (Bailey *et al*., 2017). Guidelines have been published on how both SVD and CD should be provided to pregnant women with suspected or confirmed COVID-19 in Nigeria in line with global guidance (Okunade *et al*., 2020). However, do these revamped services, which are deemed essential to be maintained even during the pandemic (WHO, 2020c), come at an additional cost to women?

The 2020 World Health Statistics Report showed that 15.1% of the Nigerian population has household expenditure on health which is more than 10% of their total household expenditure or income, a benchmark used to define catastrophic health expenditure (WHO, 2020d). This estimate is more than twice the rate in the WHO Africa region (7.3%) (WHO, 2020d). Before the advent of COVID-19, there were already established concerns with access to skilled health personnel and EmOC for the poorest women, who account for about three-quarters of maternal deaths (GBD 2015 Maternal Mortality Collaborators, 2016). Many families were shown to be shouldering maternal health service utilization costs beyond their capabilities, with trans-generational ramifications (Miller and Belizán, 2015). However, the many indirect effects of COVID-19 which had been previsioned including the disruption of health systems and the consequences of the lockdown measures implemented by many countries across the globe (Roberton *et al*., 2020), including Nigeria, brings a need to focus on the cost of utilizing essential maternity services in the middle of the pandemic. This is particularly critical for areas that have been most severely affected. Lagos is the epicentre of the COVID-19 pandemic in Nigeria with over 18,255 laboratory-confirmed and 202 deaths, compared to the national average of 1,480 cases and 28 deaths, as of 4^th^ September 2020 (NCDC, 2020a). The objective of this study is to identify and assess out-of-pocket (OOP) cost for access and utilization of maternity services for childbirth amongst pregnant women with COVID-19 in Lagos, Nigeria.

## Materials and methods

This was a hospital-based cost analysis to estimate the cost of spontaneous vaginal and caesarean deliveries, from the user’s (patient’s) perspective. The study was conducted in Lagos, a south-western state in Nigeria, which is the economic nerve centre of Nigeria and arguably the most industrialized part of the country. As the COVID-19 cases mounted in the state, two of the 24 public health facilities, the federal government-owned Lagos University Teaching Hospital (LUTH) and the state government-owned Gbagada General Hospital were tasked with providing bespoke care for pregnant women who were suspected or infected with COVID-19 (Igomu, 2020a; Ishola, 2020). While pregnant women who went to Gbagada General Hospital were not required to pay for care, as part of the Lagos State Government’s policy to cover the cost of care for all patients with COVID-19, those who used LUTH for delivery services needed to pay some cost for labour and childbirth services (James, 2020). As such, LUTH was selected as the site for this study, since it was the only facility in Lagos for which actual cost of service utilization could be captured.

LUTH is an 800 bedded tertiary referral hospital which provides health services to the over 20 million population of Lagos State (United Nations, 2019) and residents of surrounding states. Annually, the obstetrics and gynaecology department manages over 2,000 deliveries, including referrals from other secondary hospitals in the state. LUTH managed the first pregnant woman with confirmed COVID-19 in Nigeria. The hospital subsequently put together a multidisciplinary team of obstetricians, anaesthetists, neonatologists, nurses, psychiatrists and infectious disease control experts to provide care for these women (Ishola, 2020; Makwe *et al*., 2020).

For our study, the inclusion criterion was pregnant women with COVID-19 who delivered at LUTH, either by SVD or CD at term or near term. Pregnant women who delivered outside the hospital and were subsequently admitted for management of complications post-delivery, those admitted into private wards and those exempted from paying user fees were excluded. The reason for excluding these groups was that their fees to utilize care would not be the most reflective of the general population. Women were only approached after their discharge from hospital to ensure that cost of care could be fully captured. All nine pregnant women recruited for the study had laboratory-confirmed COVID-19 and were managed in LUTH between 1^st^ April 2020 and 31^st^ August 2020.

Leveraging guidance from a recently published systematic review on cost of utilising maternal health services in low- and middle-income countries (LMICs) (Banke-Thomas *et al*., 2020), we collected data on direct cost components spent within the facility (during admission), outside the facility (household), opportunity (loss of productivity) costs and any other relevant costs that women claimed to spend for their care. All of these made up total cost of service utilization. We noted any exemptions and donations that reduced the cost paid by women for service utilization.

Facility-based costs included cost paid for hospital admission, service fee, medicines, diagnostics, medical supplies and food. In capturing these costs, we separated facility-based costs related to the obstetric care of the women from those of their COVID-19 care. A detailed review of patient financial account records in the hospital was used to capture all facility-based costs. For this facility-based cost data, a verification process of double-checking was put in place to ensure there were no errors. For comparison, we collected data on the standard facility-based cost of SVD and CD for booked (registered) and un-booked (unregistered) pregnant women pre-COVID-19 as well as cost of the same services for pregnant women who presented during the COVID-19 pandemic but were not infected with COVID-19 from hospital financial records.

Household costs were those spent on transportation to and from the facility as well as the cost of childcare while on admission. Opportunity costs included loss of productivity of the caregiver who supported the woman during her admission as well as women who are self-employed and were not making any income while on admission. For this, we collected data on the monthly income of self-employed women and all the caregivers of women who were physically present to support them while on admission. In addition, we collected data on any other costs as reported by the women. In cases in which the woman could not recollect cost that was spent while on admission, she asked her husband/relatives who supported her while on admission to fill in the gaps. A pre-tested online tool was administered to women to collect the relevant household, indirect and other cost data. For this, this cost of travel was deemed to be to and from the facility. For costs related to childcare and loss of productivity, the consensus principle of allocation known as the full absorption cost principle was applied, ensuring that only the included cost was allocated to services based on actual utilization (Mogyorosy and Smith, 2005). To achieve this, we only included a pro-rated cost of the typical monthly cost related to the number of days that the women spent on admission.

All cost data were collected in local currency (Naira (N)), as per established good practice (Mogyorosy and Smith, 2005). Analysis was conducted in Microsoft Excel (Microsoft Corporation, Redmond, United States) sheet following conversion of cost data to United States Dollars (US$) as per the mean exchange rate for the year on the official OANDA Corporation website (OANDA, 2020). All costs were presented in US$, as the currency is widely understood and is the medium of exchange for many international transactions (Turner *et al*., 2019). Based on these US$ cost equivalents, the total cost of utilization of maternity services for childbirth was calculated by summing up the component costs gathered from data collection. To synthesise findings, we identified the unique obstetric and COVID-19 features of each woman which may influence the cost of care utilization. The obstetric features of interest were known pregnancy complications including abortion, ante- or post-partum haemorrhage, obstructed labour, pre-eclampsia/eclampsia, and sepsis (WHO *et al*., 2009). COVID-19 features were either mild (asymptomatic or with non-specific symptoms) or severe (with respiratory distress), as defined by the Nigeria Centre for Disease Control (NCDC) (NCDC, 2020b). Individual costs of service utilization were summed, and key cost drivers for utilization were identified for each case with care taken to recognize potential reasons for variations based on unique obstetric and COVID-19 features. We estimated the mean and median cost of the component and total costs per service (SVD, elective caesarean delivery (ELCD) and emergency caesarean delivery (EMCD)). We also estimated how much more women would have paid if there were no exemptions or donations. This allowed us to estimate the financial value of any subsidies received by the pregnant women included in our study. Where price fluctuations exist, we conducted a sensitivity analysis to test their influence on subsidy valuation. In addition, we compared the mean facility-based cost of care for pregnant women with COVID-19 with standard facility-based costs for those who presented pre-COVID-19.

Participation in the study was entirely voluntary and written informed consent was obtained from each participant who agreed to take part with no financial incentive offered. Since the study was conducted after discharge, participation or otherwise had no bearing on their care. Women were also informed that they could withdraw from the study at any time. Also, anonymity of patients was maintained in reporting study findings. Ethical approval to conduct the study was obtained from the Health Research and Ethics Committee of the Lagos University Teaching Hospital (LUTHHREC/EREV/0520/24).

## Results

### Socio-demographic and obstetric characteristics of the study participants

The age of pregnant women with COVID-19 who presented at the hospital was from 22 to 40 years, with a median age of 33 years. All nine women were married and had attained tertiary education. Six of the women were employed, one self-employed and the remaining two were unemployed. The spouses of all 9 women were gainfully employed.

Of the nine pregnant women with laboratory-confirmed COVID-19 included in the study, two remained symptomatic while on admission presenting with Acute Respiratory Distress Syndrome (ARDS), the other seven were asymptomatic until discharge. In terms of the obstetric characteristics of the included women, their expected gestational age (EGA) ranged from 33 to 40 weeks. Seven of the nine women presented with no obstetric complications during the index pregnancy, one woman presented with high blood pressure in pregnancy (preeclampsia) and another with bleeding due to abruption of the placenta. For mode of delivery, there were eight CDs (Case 1-8). Five cases were done as an elective (Case 1-5), and the other three were emergency (Cases 6-8). All CDs were done under spinal anaesthesia. Case 9 was the only patient who gave birth via SVD [Table 1].

**Table 1:** Description of care and utilization costs of spontaneous vaginal and caesarean delivery for pregnant women with COVID-19 in US Dollars

|  | Case 1 | Case 2 | Case 3 | Case 4 | Case 5 | Case 6 | Case 7 | Case 8 | Case 9 |
| --- | --- | --- | --- | --- | --- | --- | --- | --- | --- |
| **Relevant details of care** |  |  |  |  |  |  |  |  |  |
| COVID-19 symptom state | Mild | Mild | Mild | Mild | Mild | Severe | Mild | Severe | Mild |
| Obstetric complication(s) in Index pregnancy | None | None | None | None | None | None | Preeclampsia | None | Abruptio Placentae |
| Mode of delivery | ELCD | ELCD | ELCD | ELCD | ELCD | EMCD | EMCD | EMCD | SVD |
| Length of hospital stay (days) | 11 | 21 | 20 | 13 | 5 | 22 | 21 | 15 | 4 |
| **Cost of service utilisation in US\$** | | | | | | | | | |
| **Facility-based costs** |  |  |  |  |  |  |  |  |  |
| Service fee* | 0 | 0 | 0 | 0 | 0 | 0 | 0 | 0 | 0 |
| Ward admission* | 0 | 0 | 0 | 0 | 0 | 0 | 0 | 0 | 0 |
| Feeding* | 0 | 0 | 0 | 0 | 0 | 0 | 0 | 0 | 0 |
| Medicines | 202 | 331 | 248 | 246 | 244 | 594 | 210 | 235 | 22 |
| Diagnostics (obstetric) | 59 | 96 | 75 | 99 | 63 | 248 | 199 | 105 | 38 |
| Diagnostics (COVID-19) | 0 | 0 | 0 | 0 | 0 | 74 | 0 | 0 | 0 |
| Extra Oxygen consumption | 0 | 0 | 0 | 0 | 0 | 1,422 | 0 | 55 | 0 |
| Supplies/Consumables | 101 | 79 | 134 | 173 | 132 | 132 | 119 | 99 | 45 |
| Personal Protective Equipment | 478 | 597 | 446 | 445 | 303 | 463 | 229 | 274 | 116 |
| Discharge fee | 7 | 7 | 0 | 12 | 7 | 7 | 7 | 7 | 7 |
| Total facility-based costs | 847 | 1,109 | 903 | 975 | 749 | 2,939 | 764 | 773 | 228 |
| **Household costs** |  |  |  |  |  |  |  |  |  |
| Transport (To and from) | 21 | 8 | 10 | 7 | 16 | 10 | 5 | 13 | 13 |
| Childcare | 0 | 0 | 0 | 68 | 0 | 78 | 0 | 0 | 10 |
| Total household costs | 21 | 8 | 10 | 75 | 16 | 88 | 5 | 13 | 23 |
| **Other costs** |  |  |  |  |  |  |  |  |  |
| Sundry items | 0 | 0 | 0 | 0 | 52 | 0 | 0 | 0 | 0 |
| Gifts/Tips to hospital staff | 0 | 0 | 0 | 0 | 0 | 0 | 0 | 0 | 0 |
| Total other costs | 0 | 0 | 0 | 0 | 52 | 0 | 0 | 0 | 0 |
| **Opportunity costs** |  |  |  |  |  |  |  |  |  |
| Loss of productivity cost | 572 | 0 | 0 | 0 | 433 | 1,525 | 546 | 845 | 243 |
| Total opportunity costs | 572 | 0 | 0 | 0 | 433 | 1,525 | 546 | 845 | 243 |
| **TOTAL COST** | **1,439** | **1,117** | **914** | **1,049** | **1,250** | **4,553** | **1,315** | **1,631** | **494** |

*\*Patients with COVID-19 are exempted from paying service fee, ward admission and feeding.*
*EMCD = emergency caesarean delivery, ELCS =elective caesarean delivery, SVD = spontaneous vaginal delivery.*

The women spent between 4 and 22 days on admission with a median of 15 days [Table 1]. Except for one case of a macerated stillbirth, all mothers and their babies were discharged alive.

### Cost of utilising skilled birth attendance during intrapartum care

The total cost of utilization (facility-based and household) was US$494 (N190,150) for the sole pregnant woman who had SVD and mild COVID-19. Total utilization cost for those who gave birth via CD ranged from US$914 (N351,495) for a pregnant woman who had uncomplicated ELCD to US$4,553 (N1,751,165) for one who had EMCD and severe COVID-19. Mean total cost of utilization across the entire population was US$1,529 (N588,765) with a standard deviation of US$1,112 (N428,246). When disaggregated, facility-based costs made up the highest proportion of utilization costs for all the women (67% of the mean total cost of utilization) while opportunity cost due to loss of productivity of the caregiver for the woman while on admission made up 30% [Table 1]. Transport, childcare and purchase of other sundry items constituted the remaining 3% of the total cost of utilization. For transport, three of the women (Case 2, 4 and 7) used ambulance services offered by the Lagos State COVID-19 response team to reach LUTH, at no cost to them. However, travel from the hospital back to their places of residence cost them between US$5 (N2,000) and US$8 (N3,000). Childcare came at no cost to six of the nine women, as they had relatives who minded their children while they were on admission [Table 1].

For facility-based costs, the hospital management exempted all COVID-19 patients from paying the service fee, ward admission and feeding, in line with the Federal Government’s directive for the management of COVID-19. In addition, laboratory confirmation for COVID-19 by Reverse Transcription Polymerase Chain Reaction (RT-PCR) test was free, including the serial testing required by the patient until they were confirmed negative. Furthermore, with support from the Federal Government, international agencies, some Non-Governmental Organizations (NGOs), charities, and philanthropic personnel, some personnel protective equipment (PPE) including N95 masks, protective gowns, gloves, face shields and aprons were made available to skilled health personnel to support safe provision of care, at no cost to the women.

For the costs still required, the woman who had SVD paid a total of US$228 (N87,750). Cost of additional personal protective equipment (PPE) required for their care was the major cost driver (50%). This was followed by supplies (20%) and obstetric diagnostics (17%). For ELCD, facility-based cost ranged from US$749 (N287,950) to US$1,109 (N426,685), with a median cost of US$903 (N347,495). Major cost drivers for ELCD were PPE (50%), medicines (28%), and medical supplies (14%). Excluding the cost of additional supplemental oxygen required by women who had severe COVID-19 symptoms, EMCD cost from US$719 (N276,445) to US$1,517 (N583,478). The major cost drivers were medicines (35%), PPE (32%), and diagnostics (18%). Based on severity of COVID-19 symptoms, cost ranged from US$228 (N87,750) for a woman with mild disease who was on admission for four days and gave birth via SVD to US$2,939 (N1,130,498) paid by a woman who had severe COVID-19 symptoms requiring additional supplemental oxygen extra-operatively while on admission. For this latter case (Case 7), the woman also had to pay for additional investigations required to manage the ARDS, including chest x-ray and arterial blood gases. For this case, medical oxygen required to manage severe COVID-19 symptoms was the major cost driver (48%), followed by medicines (20%), and medical supplies (14%) [Table 1].

Despite the cost exemptions, there are significant cost differences between facility-based cost paid by pregnant women with COVID-19 and those paid in the pre-COVID-19 era when all pregnant women were required to pay a service fee and ward admission. During the pandemic, the cost of SVD has more than doubled the cost paid by a booked pregnant woman pre-COVID (US$113 (N43,400). For CD, excluding medical oxygen, the average facility-based cost of all eight CD patients (US$984 (N364,551)) is about 2.5 times more than what women paid pre-COVID (US$384 (N147,660)) [Table 1 and Table 2].

**Table 2:** Facility based cost of utilizing spontaneous vaginal and caesarean delivery pre-COVID-19 in US Dollars

|  | SVD - Booked | SVD - Un-booked | CS (Spinal anaesthesia)<br>- Booked | CS (General<br>anaesthesia) - Booked | CS (Spinal anaesthesia)<br>– Un-booked | CS (General anaesthesia)<br>– Un-booked |
| --- | --- | --- | --- | --- | --- | --- |
| **Facility-based costs in US\$** | | | | | | |
| Service fee* | 28 | 55 | 82 | 82 | 82 | 82 |
| Ward admission | 0 | 0 | 74 | 74 | 74 | 74 |
| Feeding | 0 | 0 | 0 | 0 | 0 | 0 |
| Medicines | 42 | 42 | 148 | 175 | 148 | 175 |
| Diagnostics (obstetric) | 10 | 75 | 74 | 74 | 126 | 126 |
| Antenatal fees | 28 | 0 | 0 | 0 | 0 | 0 |
| Supplies/Consumables | 0 | 0 | 0 | 0 | 0 | 0 |
| Discharge fee | 7 | 7 | 7 | 7 | 7 | 7 |
| **Total facility-based costs** | **113** | **179** | **384** | **411** | **436** | **464** |

*\*Service fees paid for vaginal delivery include ward admission and feeding.*

If there were no exemptions and donations, the pregnant woman with mild COVID-19 who gave birth via SVD (Case 9) would have paid US$526 (N202,250) as facility-based costs, meaning she received 57% of the facility-based cost as subsidies and donations. Pregnant women with mild COVID-19 requiring CD (Case 1-5 and 7) would have paid between US$1,767 (N679,470) and US$1,960 (N754,005), though their actual costs were subsidised by between 43% and 66%. Those with severe COVID-19 symptoms requiring CD would have paid between US$2,181 (N839,165) and US$5,088 (N1,957,118), though their actual costs were subsidised by between 42% and 65% [Table 3]. Using the most conservative estimates on potential cost subsidies being received by the women, facility-based costs were subsidised by between 21% and 51% [see <u>Supplementary Data</u>].

**Table 3:** Subsidies received by women due to donations and exemptions in US Dollars

| Case | Days in Delivery Theatre | Delivery and immediate post-partum (\$) | First day post-partum till discharge (\$) | Hospital admission fees (\$) | Additional days beyond week 1 (\$) | Total subsidy received (\$) | Actual amount paid by women (\$) | PPE cost less cost paid by women for PPE (\$) | Total that would have been paid (\$) | % paid by women | % paid by other sources |
| --- | --- | --- | --- | --- | --- | --- | --- | --- | --- | --- | --- |
| 1 | 8 | 780 | 770 | 74 | 36 | 1,264 | 847 | 478 | 2,111 | 40.11% | 59.89% |
| 2 | 4 | 780 | 385 | 74 | 127 | 851 | 1,109 | 597 | 1,960 | 56.59% | 43.41% |
| 3 | 4 | 780 | 385 | 74 | 118 | 993 | 903 | 446 | 1,896 | 47.64% | 52.36% |
| 4 | 4 | 780 | 385 | 74 | 55 | 930 | 975 | 445 | 1,905 | 51.17% | 48.83% |
| 8 | 4 | 780 | 385 | 74 | 0 | 1,018 | 749 | 303 | 1,767 | 42.38% | 57.62% |
| 5 | 16 | 780 | 1,539 | 74 | 137 | 2,149 | 1,517 | 463 | 3,666 | 41.38% | 58.62% |
| 6 | 7 | 780 | 673 | 74 | 127 | 1,508 | 764 | 229 | 2,272 | 33.62% | 66.38% |
| 7 | 7 | 780 | 673 | 74 | 73 | 1,408 | 719 | 274 | 2,127 | 33.79% | 66.21% |
| 9 | 2 | 127 | 71 | 28 | 0 | 110 | 228 | 116 | 338 | 67.54% | 32.46% |

[Insert Supplementary Data: Scenario testing for exemptions and donations influencing out-of-pocket costs]

## Discussion

This study was conducted to identify and assess the out-of-pocket cost of childbirth services for women in Nigeria during the COVID-19 pandemic. Our findings suggest that pregnant women with COVID-19 are paying significantly higher fees for SVD and CD, despite cost exemptions on certain cost components that have been instituted. The major driver in this increased cost of utilization relates to the PPEs required for care of patients infected with mild COVID-19. Medical oxygen is the major cost driver when pregnant women present with severe COVID-19 and require long admission in the hospital.

Regarding facility-based cost of utilising care, we found that pregnant women with COVID-19 are paying as much as US$228 (N87,750) for SVD when they have mild COVID-19 and US$2,939 (N1,130,498) for EMCD when they present with severe COVID-19. In a 2020 systematic review, authors reported that the median cost of utilising SVD across LMICs was US$40 in a public hospital while CD was US$178 in public hospitals (Banke-Thomas *et al*., 2020). Our findings suggest that COVID-19 pregnant women are paying six times more for SVD and as much as 16 times more if they have severe COVID-19 and require CD. As per the same 2020 systematic review (Banke-Thomas *et al*., 2020), the facility-based cost being estimated for SVD is almost equal to the highest cost reported for private hospital SVD use in Nepal of US$295 (Acharya *et al*., 2016). Similarly, our mean facility-based cost for CD for pregnant women with COVID-19 of US$1,132 is over twice the highest cost for the same service in private hospitals in India (US$497) after transport and opportunity costs have been excluded (Banke-Thomas *et al*., 2020). This finding is particularly concerning, especially as most women receive care from public hospitals, which are viewed as a cornerstone for achieving universal health coverage in LMICs (Sachs, 2012).

It is established that tertiary hospitals like LUTH are significantly more expensive for care compared to secondary and primary facilities, mostly because of the specialist expertise that is concentrated therein (Banke-Thomas *et al*., 2020). However, it is important to keep in mind that the standard cost for an un-booked patient managed in LUTH pre-COVID (US$464 [Table 2]) is still less than the maximum obtainable cost reported for another Nigerian teaching hospital (US$667) in 2013, as per published peer-reviewed literature (Adamu *et al*., 2013), though it is deemed exorbitant by many Nigerian women (Wright *et al*., 2017). In our study, pregnant women with COVID-19 are paying as much as two times more for SVD and three times more facility-based costs for CD when compared to the pre-COVID era. This is despite the government-mandated cost exemptions on certain cost components that have been instituted and the donations that have been received to support care provision (Alagboso and Abubakar, 2020; Igomu, 2020b; Naeche, 2020). The major driver in this increased cost of utilization for SVD and ELCD relates to the PPEs required for care of patients infected with COVID-19. Yet, our findings show that pregnant women are not even being required to pay for all the PPE that is used to support their care. This emergence of PPEs as major cost drivers (as much as 50%) is concerning. Previously, though cost drivers varied by country, most reported that medicines and supplies, transport, and lodging were the major cost drivers that women had to tackle to access care (Banke-Thomas *et al*., 2020). However, there is also the emergence of medical oxygen as the major cost driver (as much as 48%) in the severe cases that require long hospital admission. This is despite the fact that it is the second most important component of the COVID-19 care package (Stein *et al*., 2020).

Informal payments in the form of gifts and tips to health workers have been reported in several studies conducted in other LMICs (Afsana, 2004; Borghi *et al*., 2006; Kruk *et al*., 2008; Khan and Zaman, 2010). However, in our study, no woman reported giving any gifts to health workers. With so much concern and caution being taken with care of pregnant women with COVID-19, it might be the case that the women are simply not giving gifts. However, this is unlikely, as pregnant women in Nigerian have been described as usually being very appreciative of the efforts of health workers in taking care of them (Adamu *et al*., 2013). A more plausible explanation may be that the health workers themselves were refusing to receive gifts or tips from women or their possibly asymptomatic relatives because they wanted to minimize contact, conscious of the possibility of being infected through the gifting.

For the other cost components, median transport costs to and from the hospital of US$10 reported in our study is higher than has been reported in Tanzania (US$0.09) but significantly lower than US$51 reported in Bangladesh (Afsana, 2004; Kruk *et al*., 2008). Estimates of the loss of productivity for time spent by the caregiver in our study were significantly higher than what has been reported in the literature before COVID. In our study, opportunity costs ranged from US$243 to US$572, while in the literature, adjusted estimates ranging from US$3 in Lao PDR for SVD (Douangvichit *et al*., 2012) to US$89 for caesarean delivery in Nepal have been reported (Acharya, 2016). It is difficult to explain this finding. It might be the case that the partners in our study make greater losses for not working while providing care, because their jobs are more lucrative than the Lao PDR and Nepal study. Another plausible reason may be that the pregnant women with COVID-19 in our study had to stay longer on admission, and as such, their partners had to stay longer away from work.

There are clear policy implications of our study findings. Pre-COVID-19, cost of utilising delivery services was already deemed as exorbitant for women. In a 2013 Nigerian study, the authors found that the mean total expenditure for childbirth including facility-based and household costs (US$246.30 (N39,400)) was more than the monthly family income for 94.6% of respondents (Adamu *et al*., 2013). However, our study now shows that the financial burden is even significantly higher for pregnant women with COVID-19. It is particularly important to highlight that service fees have been excluded for pregnant women with COVID-19. However, such exemptions sometimes do not go far enough (Kruk *et al*., 2008), especially if they are going to be replaced by new costs like PPE and medical oxygen.

Pre-COVID-19, health care providers had some form of PPE to protect them when providing services, albeit not as many as is required for adequate and safe care today. Indeed, demand now by far outstrips supply globally (Burki, 2020), with 60% of health care workers reporting that they have not had access to sufficient PPE to keep them safe while providing care to pregnant women (Semaan *et al*., 2020). With such gaps in the PPE supply chain, costs are passed to women as shown in our study. This increases the risk of catastrophic health expenditure. Providers, more so those in LMICs, who are facing greater shortages and had huge budget constraints even pre-COVID, need to explore more innovative ways to source PPEs without passing the burden unto pregnant women (Green *et al*., 2020). Indeed, at the beginning of the pandemic, the cost of PPEs was cheaper, but this has now skyrocketed with the price of surgical masks having increased sixfold, N95 respirators by threefold, and surgical gowns have doubled (Burki, 2020). While government-led strategies such as loosening import regulations and commandeering business to accelerate supply have been recommended (Livingston *et al*., 2020), there is a case for the government to scale up local manufacturers to explore PPE production at reduced rates (Olatunbosun, 2020). Also, negotiations with sellers, while offering incentives for reduced costs and regulation of sell-on costs will help to drive down the cost and reduce excessive mark-ups being passed on to pregnant women. As a strategy for preparing for future pandemics, it is important to use data to work out actual PPE needs and stockpile PPEs, minimizing the risk of costs being transferred to women.

New thinking is also needed for the supply of medical oxygen. When this cost is added on to the facility-based obstetric cost, they can double the total facility-based cost of utilization, especially with long-term admission. Before the COVID-19 pandemic, there was already concern about oxygen sufficiency in many African countries (Howie *et al*., 2009; Aluvaala *et al*., 2015; Dauncey *et al*., 2019). While innovative approaches such as installing oxygen concentrators as opposed to the more expensive cylinders, enabling private construction of oxygen plants within hospitals, and use of solar-powered oxygen delivery are being implemented to boost oxygen supply in some African countries (Stein *et al*., 2020), it is important that these costs are not passed on to pregnant women.

In the pre-COVID era, there was mixed evidence on the effectiveness of user fee removal. While some suggest it leads to increase in utilization of facilities for delivery (Witter *et al*., 2010), others have submitted that it can increase the workload for the already overworked health workforce and result in supply gaps of medicines and supplies for pregnant women (Gilson and McIntyre, 2005). However, these are uniquely challenging times when reduced economic activity and the need for stay at home with children who are not able to go to school means many women and their partners may not be able to afford user fees that they would have normally been able to afford or at least sourced from somewhere. It should be noted that the women in our study were all educated and they and/or their partners were employed, yet, as our results showed, they benefited between 32 and 62% of subsidies in facility-based costs. With 40% of the population living below the poverty line (US$360 (N137,400) per year) (Varrella, 2020), many will not be able to afford these increased service utilisation of the COVID-19 era, without these donations and exemptions (Alagboso and Abubakar, 2020; Igomu, 2020b; Naeche, 2020). Indeed, there might be a case for a comprehensive fee exemption policy, as was done by the state government in Gbagada General Hospital, where pregnant women did not have to pay a cent in facility-based costs (James, 2020). However, it is not known how long this can be sustained, with the government spending as much as (US$260 (N100,000)) and (US$2,604 (N1,000,000)) per day on off-setting bills of patients with COVID-19 (Adediran, 2020). Similarly, how long can donations last?

Another key issue lies within the community, where there is a real fear of contracting COVID-19 in the hospital and as such women do not actually want to have their babies in the hospital (Semaan *et al*., 2020). There are also curfews, lockdowns, and transport restrictions which women need to navigate to reach health services (Oneko, 2020; Semaan *et al*., 2020). Adding a financial barrier to this would only further disenfranchise women and allow them to even more rationally explain why they do not want to access skilled care in a facility, COVID-19 positive or otherwise. In any case, if COVID-19 truly becomes the “new normal” as is often said, costs of maternity services for childbirth may yet still go up for all pregnant women with some experts already proposing the need for universal testing of pregnant women for COVID-19 and a lower threshold for admitting pregnant women to hospital and intensive care unit (Breslin *et al*., 2020; Silveira Campos and Caldas, 2020). This and any other additional costs may cause pregnant women to delay care-seeking, putting them at a greater risk of otherwise preventable obstetric complications (Banke-Thomas *et al*., 2017; Kyei-Nimakoh *et al*., 2017). Indeed, as these costs still need to be paid, the COVID-19 pandemic might be the perfect opportunity to drive advocacy for more enrolment in the National Health Insurance Scheme, which has only 2.1% of women of reproductive age group in Nigeria registered on it (Aregbeshola and Khan, 2018).

With such prohibitive costs associated with care of pregnant women with COVID-19, our findings suggest that the age-old adage “prevention is better than cure” might have substantial cost savings implication for pregnant women and indeed for LMIC governments who are still choosing to bear the cost of service utilization. While current guidance proposes telemedicine as a platform to provide ante-natal care (ANC) for pregnant women with suspected or confirmed COVID-19 (Okunade *et al*., 2020), scaling this up to include all pregnant women requiring ANC while counselling them to minimize risk of infection around them might help forestall additional cost required to manage COVID-19 in pregnancy. However, this should not be applied as a one-size-fits-all solution, especially for high-risk pregnancies, who need to be seen physically, and those who do not have ‘smart’ phones’ enabled with telemedicine capacity (Green *et al*., 2020).

Our study has some key strengths. Firstly, this is the first paper that reports the actual cost of delivery service utilization in our paper in the middle of the ongoing pandemic. To do this, we captured primary cost data, and leveraged the best available methodological guidance on conducting costing studies and insights from a recently published systematic review (Mogyorosy and Smith, 2005; Banke-Thomas *et al*., 2020). Our study is also a whole population sample, capturing all nine pregnant women who have presented with COVID-19 in the largest teaching hospital of the epicentre of COVID-19 in Nigeria. It also had enough heterogeneity to allow a broad understanding of the financial burden faced by women. In addition, we have leveraged multiple data sources to capture a full economic cost implication of service utilization. However, there are limitations to bear in mind in interpreting findings of this study. Firstly, our sample size is small, with only one SVD. Though this is not dissimilar to other studies with a recent systematic review reporting 92 women across nine studies which reported an 80% CD rate (Smith *et al*., 2020). One other limitation relates to the recall bias that our findings may be subject to, as we captured household costs entirely based on what they could recollect following their discharge. However, having to double-check with their partners regarding the cost data being provided helped to minimize any influence of this bias. In addition, we have only reported cost from one public tertiary hospital in Nigeria, and this cost may not be representative of the cost being incurred by women around the country, especially within the private sector, where costs for using services are typically higher than in the public sector (Banke-Thomas *et al*., 2020). In addition, we did not collect or report household costs data in the pre-COVID era. However, the cost of many products and services, including transportation has gone up in this period (Mogaji, 2020). As such, this additional data would not have altered our conclusion that women are paying more for childbirth during the COVID-19 pandemic.

## Conclusion

In many LMICs, including Nigeria, hard-won gains were already being made with reducing catastrophic health expenditure for pregnant women, realising universal health coverage and improving maternal and newborn health outcomes. However, COVID-19 has disrupted the normal, necessitating new thinking to protect these gains (Graham *et al*., 2020). Cost of utilizing maternity services for childbirth have increased and are likely to remain significantly high for women if exemptions being offered by governments become unaffordable, donations reduce or new requirements for universal testing have a chargeable fee. If COVID-19 becomes the new normal, then there will be many more pregnant women with COVID-19, including many who cannot afford the huge costs of care. Urgent measures are needed to ensure that women and their families are not being locked out of the health system.

## Data Availability

All data referred to in the manuscript are presented in the manuscript.

## ACKNOWLEDEGEMENTS

We are indebted to all the staff of the Lagos University Teaching Hospital who made data collection for this study possible and would like to particularly acknowledge the support of Pharmacist Olabisi Opanuga. We are especially grateful to the pregnant women with COVID-19 who took out time post-discharge to take part in this study.

